# Recognition of Diabetes in a Multi-ethnic Population: Results from a Nationally Representative Population-Based Study

**DOI:** 10.1101/2021.02.01.21250802

**Authors:** Kumarasan Roystonn, Jue Hua Lau, PV AshaRani, Fiona Devi Siva Kumar, Peizhi Wang, Sum Chee Fang, Eng Sing Lee, Chong Siow Ann, Mythily Subramaniam

**Affiliations:** Research Division, Institute of Mental Health, 10 Buangkok View, Singapore 539747; Clinical Research Unit, Diabetes Centre, Admiralty Medical Centre, Singapore 730676; National Healthcare Group Polyclinics, 3 Fusionopolis Link. Nexus@One-North, 06-13, Singapore 138543; Saw Swee Hock School of Public Health, National University of Singapore, Singapore 117549

## Abstract

**Background:** The prevalence of diabetes is on the rise in developed countries. Yet discrepancies exist in reports regarding the level of knowledge of diabetes. This study evaluates the recognition of diabetes among residents in the Singapore population. Sociodemographic characteristics associated with the ability to correctly recognise diabetes were also examined.

**Methods:** This was a nationwide cross-sectional vignette-based study. Data were collected from 2895 residents aged 18 years and above through face-to-face interviews, of which 436 are persons with diabetes. Descriptive statistics, chi-square tests, and binary logistic regression were used in the analyses.

**Results:** In total, 82.7% (2418/2895) of respondents recognised diabetes correctly. In adjusted models, correct recognition was significantly higher among respondents aged 35-49 years (OR, 1.85; 95% CI, 1.15 to 2.98; P= 0.01), 50-64 years (odds ratio [OR], 2.06; 95% confidence interval [CI], 1.19 to 3.56; P= 0.01), ethnic Malays (OR, 1.39; 95% CI, 1.02 to 1.89; P= 0.04) (vs. Chinese) and persons with diagnosed diabetes (OR, 2.64; 95% CI, 1.38 to 5.08; P < 0.01). Being male (OR, 0.64; 95% CI, 0.46 to 0.90; P < 0.01), ethnic Others (OR, 0.59; 95% CI, 0.37 to 0.93; P < 0.01), and being unemployed (OR, 0.48; 95% CI, 0.25 to 0.92; P= 0.03), were significantly associated with poor recognition of diabetes.

**Conclusion:** Overall public recognition of diabetes is high, but the significant gaps in knowledge in certain demographic groups are of concern. Public health interventions aimed at preventing and controlling diabetes should continue to target all members of the population with accurate and appropriate information. Ongoing efforts of diabetes awareness and educational programs need to be improved, particularly for young adults, males, and the unemployed.

## INTRODUCTION

Diabetes mellitus is a prevalent chronic condition which results in substantial target organ disease and significant complications including blindness, kidney failure, stroke and coronary heart disease. In 2017, it was estimated that there were 451 million adults (8.5%) worldwide with diabetes, with an associated cost of US$850 billion comprising direct and indirect medical costs as well as informal care.[1, 2] There is a growing diabetes epidemic in the Asia-Pacific region with more than 50% of people with diabetes being undiagnosed.[2, 3] To date, the International Diabetes Federation (IDF) has estimated that 88 million adults live with diabetes in this region with a projected increase to 153 million by 2045.[3] In Singapore, as in many developed countries, diabetes is a major public health concern.[4] The cost of diabetes per patient was found to be US$1,575.6, which was higher than the costs reported in other Asian countries.[5] In 2017, diabetes became the seventh leading cause of morbidity and premature death in Singapore.[6] An epidemiological study of the resident general population, found the prevalence of diabetes to be approximately 11.3% (one in nine) in those aged 18 years and above with higher rates among men than women[7]. The national survey data also revealed that 51.4% of individuals in Singapore were unaware that they had diabetes.

Diabetes literacy is herein defined as the knowledge and beliefs regarding diabetes mellitus that aid recognition, management, or prevention of this physical disease. This is adapted from ‘mental health literacy’, a term by Jorm[8] with widespread usage that refers to “knowledge and beliefs about mental disorders which aid recognition, management, or prevention”. Poor knowledge of the signs and symptoms of diabetes may be a particularly important barrier to help-seeking behaviour for this chronic disease. Failure to recognise signs and symptoms associated with diabetes could lead to delays in timely medical attention.[9] Accurate recognition and labelling of the disease are paramount to the prevention of diabetes to facilitate early and appropriate help-seeking to improve long-term health outcomes.

Recent qualitative studies exploring health literacy in diabetes in Asian cultures found that culture shaped understanding and experiences of health literacy.[10, 11, 12]. There are significant ethnic differences in how physical health conditions are recognised and labelled. For example, in a study with ethnic Chinese, the researcher found that a majority held on to cultural beliefs that ageing or a ‘weak body’ was the cause for diabetes.[13] Much of the available literature tends to focus on the knowledge of patients with diabetes. Few reports are available on evaluating diabetes literacy in the general population, even though diabetes is increasingly becoming a major threat to global public health. A general population study of Australian adult residents found that only 14% - 29% of the population were able to correctly recognise diabetes symptoms and warning signs.[14] In the United Kingdom, 59.4% of the public were able to correctly identify diabetes symptoms and deemed to have adequate awareness of the disease.[15] On the other hand, research conducted in Sri Lanka reported that 77% of the general public surveyed could correctly recognise diabetes.[16] In Singapore, the only local study evaluating general public’s knowledge of diabetes reported that about 60% were able to recognise the symptoms, and complications of the disease.[17] However, recent literature remains sparse and studies were often with smaller samples and the findings are not a valid representation of the overall population of the country.

Singapore is a multicultural, multi-ethnic and multilingual country where the major ethnic groups of Chinese, Malays, and Indians still maintain somewhat separate cultural identities. The population of Singapore comprises about 5.9 million people, of which 3.9 million are Singapore citizens or permanent residents. The resident population comprises four main ethnic groups -Chinese (74.3 %), Malay (13.3 %), Indian (9.1 %), and Others (3.3 %).[18] Therefore, Singapore is an ideal location to study diabetes literacy among the multi-ethnic Asian population. The purpose of this population-based diabetes literacy study was to specifically evaluate the proportion of the Singapore public who are able to recognise diabetes correctly and the factors associated with it, in a nationally representative multi-ethnic sample. The results of the study may have strategic implications for the reduction of ethnic or socioeconomic disparities in diabetes detection, and better inform future health promotion campaigns.

## METHODS

### Setting and study design

This nationwide cross-sectional study of the Singapore population includes citizens and permanent residents aged 18 and above, belonging to the four major ethnic groups, who were literate in English, Chinese, Malay or Tamil and living in Singapore at the time of survey. All residents who were uncontactable due to incomplete or incorrect addresses and those living outside of the country were excluded from the study. The overall response rate of the study is 66.2%. The study was approved by the Institutional Research Review Committee and the National Healthcare Group Domain Specific Review Board (Ref no. 2018/00430). Written informed consent was obtained from all respondents 21 years of age and above, as well as from parents or guardians of those aged 18 to 20 years.

### Sample size calculation and sampling

Statistical power calculations for binary proportions post-adjusted for design effects determined sample sizes for population prevalence estimate, as well as for subgroup (age and ethnicity) estimates, with overall precision of 2.5%.[19] Using 20% as a prevalence estimate based on previously reported prevalence rates of diabetes knowledge in Singapore,[17] a total sample size of 3000 was estimated to be adequate to determine the general knowledge of diabetes in the population. The margin of error for the overall prevalence estimate was found to be 2.5%, while that of the subgroups by age and ethnicity ranged from 4.5% to 5%. The relative standard error (RSE) was found to be substantially below the acceptable range (<30%), ranging from 2.1% to 4.2%. Further details of the sampling strategy and processes are published elsewhere.[19]

The sampling frame was an administrative database of all residents in Singapore from which the sample was derived. A disproportionate stratified sampling design by age and ethnic groups was utilised in the study to randomly select a probability sample, based on 12 strata according to ethnicity (Chinese, Indian, Malay) and age groups (18-34, 35-49, 50-64, 65 & above). The study oversampled those of Malay and Indian ethnicities, as well as residents aged 65 and above in order to ensure sufficient sample size and to improve the reliability of parameter estimates for these population subgroups.

### Data collection

Data were collected in face-to-face interviews by trained interviewers, using the computer assisted personal interviews (CAPI) on handheld tablets. For quality assurance, at least 10 % of the completed interviews were validated through face-to-face and telephone follow up.

### Questionnaire

A structured questionnaire was used to obtain sociodemographic information including age, gender, ethnicity, marital status, personal income, educational and employment status, and self-reported diagnosis of diabetes. The study was introduced as an investigation of Singaporeans’ knowledge, attitudes and practices of a ‘chronic physical condition’. This was done so as to not influence respondents’ responses to the vignette presented at the beginning of the interview. The actual disease of interest, diabetes, was only revealed after the vignette section.

### Vignette adaptation

Respondents were presented with a hypothetical vignette describing a person with diabetes mellitus. The vignette was developed and refined by the researchers in consultation with experienced clinicians specialising in diabetes care. The vignette length was approximately 110-120 words and described classic and common symptoms of the disease. Presented in English, Chinese, Malay, or Tamil, the vignette was phrased with simple laymen’s terms. Further incorporating elements of the local context such as descriptions of the character’s background and home functioning, facilitated the development of a vignette storyline that was natural and relatable.[20, 21] The vignette also described a person of the same gender and ethnicity as the respondent. For instance, a Chinese male participant was presented a vignette about Mr. Tan (see Appendix A). Respondents were asked what they thought the person described in the vignette was suffering from and to name the condition associated with the vignette description (free response). They were asked to base their considerations on the available information only. The response was coded as correct if the participant was able to correctly label the condition. In cases where the response was a near approximation of the correct answer, three of the investigators including the first author (AP, KR, MS) would come to a consensus on how that response should be coded.

### Vignette translation and cognitive testing

The translation procedure undertaken was aimed at achieving conceptual equivalence using an adapted four-step process from WHO: (1) forward translation: the vignette and follow up questions were translated into the three local languages – Chinese, Malay and Tamil using a professional translating firm, (2) expert panel review: which involved a critical evaluation of expert panel recommendations to issues identified with translations (3) pre-testing and cognitive interviews (CI): further CI were undertaken, with 25 respondents from diverse age groups, ethnicity, gender, and socioeconomic (or education attainment) status. Trained cognitive interviewers systematically probed respondents on what they thought the vignette was about, what came to their mind when they were presented a particular phrase or term and were asked how they decided on their response. Words or expressions that were not easily understood, or deemed offensive or unacceptable were highlighted to interviewers, and where alternative words or expressions existed, respondents were asked which of the alternatives better conforms to their usual language. (4) The development of final translations was achieved after minor changes based on the information gathered from the CI.

### Statistical analyses

The survey sample was weighted by age and ethnicity to match the Singaporean resident population so that the results could be generalised to the population. Weighted mean and standard error of the mean were calculated for continuous variables, and frequencies and percentages for categorical variables. Descriptive statistics were performed to establish the prevalence of diabetes literacy as well as to describe sociodemographic characteristics of the study sample. Univariate analyses (t-test or Chi-square test) was used to investigate differences among age groups (18–34=1; 35–49=2; 50–64=3; 65 & above=4), gender (female=1; male=2), ethnicity (Chinese=1; Malay=2; Indian=3; Others=4), education (degree & above=1; primary & below=2; secondary=3; pre-university/junior college=4; vocational institute/ITE=5; diploma=6), employment (employed=1; economically inactive*=2; unemployed=3), income (in SGD) (below 2,000=1; 2,000 to 5,999=2; 6,000 & above=3), and diabetes diagnosis (no=1; yes=2). Categories coded as 1 were set as the reference category for all variables. A logistic regression analysis using survey weights to account for complex survey design was conducted to determine the sociodemographic variables significantly associated with correct recognition of diabetes. The level of statistical significance was set at P <0.05 using two-sided tests.

## RESULTS

The demographics of this study population are shown in Table 1. The survey data include 2895 respondents. The mean age of respondents was 45.8 years and 51.6% of the respondents were female. Majority (75.8%) were Chinese, 12.7% were Malays, 8.6% were Indians, and 2.9% were from the ethnic group, Others. For education level, 29.5% of the respondents were university graduates, 20.3% of the respondents had completed secondary education, and 20.4% had primary education or less.

**Table 1.** Sample characteristics (n= 2895)

|  |  |  | Non-diabetic<br>(n = 2,459) |  | Diabetic<br>(n = 436) |  |
| --- | --- | --- | --- | --- | --- | --- |
|  | N | Weighted | N | Weighted | N | Weighted |
| **Age group** |  |  |  |  |  |  |
| 18 to 34 | 823 | 29.9% | 817 | 32.7% | 6 | 1.8% |
| 35 to 49 | 719 | 28.2% | 670 | 29.8% | 49 | 12.3% |
| 50 to 64 | 774 | 26.8% | 591 | 24.5% | 183 | 49.1% |
| 65 and above | 579 | 15.1% | 381 | 13.0% | 198 | 36.9% |
| **Gender** |  |  |  |  |  |  |
| Female | 1,474 | 51.6% | 1258 | 52.2% | 216 | 44.9% |
| Male | 1,421 | 48.5% | 1201 | 47.8% | 220 | 55.2% |
| **Ethnicity** |  |  |  |  |  |  |
| Chinese | 796 | 75.8% | 731 | 76.9% | 65 | 64.9% |
| Malay | 974 | 12.7% | 811 | 12.1% | 163 | 18.7% |
| Indian | 918 | 8.6% | 725 | 7.9% | 193 | 15.1% |
| Others | 207 | 2.9% | 192 | 3.0% | 15 | 1.4% |
| **Education** |  |  |  |  |  |  |
| Primary and Below | 637 | 20.4% | 456 | 18.3% | 181 | 40.8% |
| Secondary School | 684 | 20.3% | 552 | 19.7% | 132 | 26.6% |
| Pre-U/Junior College | 126 | 4.8% | 112 | 5.0% | 14 | 2.3% |
| Vocational Institute/ITE | 267 | 6.6% | 241 | 6.8% | 26 | 5.2% |
| Diploma | 479 | 18.5% | 442 | 19.0% | 37 | 12.8% |
| Degree and above | 702 | 29.5% | 656 | 31.2% | 46 | 12.5% |
| **Marital status** |  |  |  |  |  |  |
| Married/cohabiting | 1,860 | 61.7% | 1,531 | 60.0% | 329 | 78.1% |
| Single | 731 | 29.2% | 704 | 31.7% | 27 | 4.7% |
| Divorced/separated | 154 | 5.0% | 131 | 5.0% | 23 | 4.9% |
| Widowed | 149 | 4.1% | 92 | 3.3% | 57 | 12.3% |
| **Employment** |  |  |  |  |  |  |
| Employed | 1,933 | 70.5% | 1,731 | 72.4% | 202 | 51.1% |
| Economically inactive<sup>a</sup> | 829 | 25.4% | 617 | 23.7% | 212 | 41.8% |
| Unemployed | 133 | 4.1% | 111 | 3.8% | 22 | 7.1% |

<sup>a</sup>Economically inactive includes retired, homemaker, student, and the physically disabled.

**Table 1. Sample characteristics (n= 2895) cont'd.**
| **Monthly Income (SGD)** |  |  |  |  |  |  |
| --- | --- | --- | --- | --- | --- | --- |
| Below 2,000 | 1,455 | 45.3% | 1,155 | 43.3% | 300 | 65.0% |
| 2,000 to 3,999 | 698 | 23.9% | 627 | 24.6% | 71 | 17.3% |
| 4,000 to 5,999 | 318 | 12.8% | 295 | 13.2% | 23 | 8.3% |
| 6,000 to 9,999 | 183 | 7.8% | 167 | 8.2% | 16 | 4.0% |
| 10,000 & above | 117 | 5.7% | 104 | 5.9% | 13 | 3.3% |
| Undisclosed | 124 | 4.5% | 111 | 4.7% | 13 | 2.1% |

Note: Frequencies and percentages may not tally to 100% due to missing data; SGD: Singapore Dollar

Table 2 presents the percentage of respondents endorsing each category with respect to recognition of the vignette. In total, 82.7% (n=2418) of the respondents correctly identified the disease from the vignette, while 0.7% (n=23) were found to have partly correct recognition (e.g., mislabelling and referring to diabetes as “high blood sugar”). The 23 responses were as such included under “correct recognition” in regression analysis. About 7.7% (n=220) incorrectly recognised the condition as other medical problems, 1.0% (n=33) of the respondents mislabelled the disease as a non-medical problem, and 4.9% (n=126) did not provide answers, responding with “don’t know” or “not sure”.

**Table 2.**
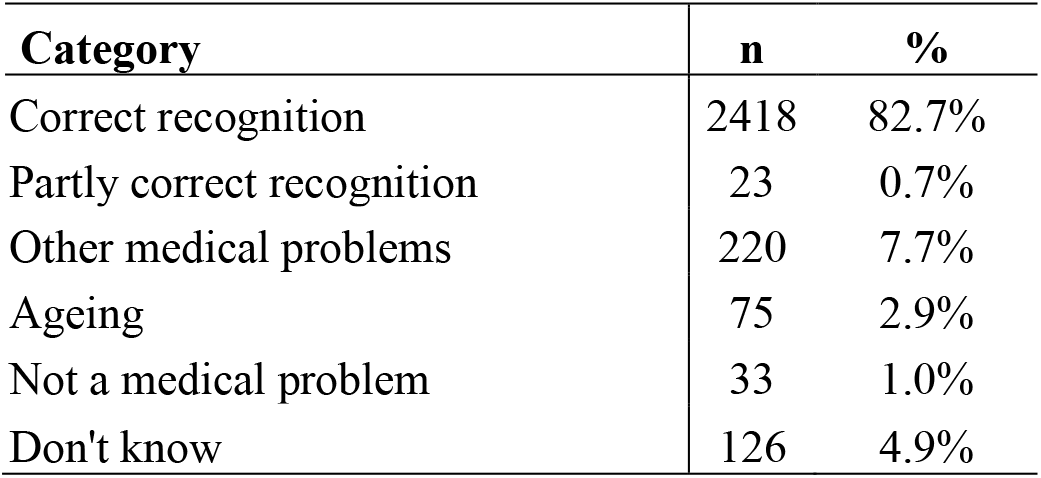
Percentage of respondents by categories of recognition of the vignette (n= 2895)

Among the 454 respondents who incorrectly recognised diabetes, the majority were male (56.1%, n=262), aged 18-34 years (40.4%, n=172), married (54.4%, n=253) and an equal proportion of them had primary education and below (22.8%, n=119), or had attained a university degree and above (22.8%, n=86). About 4.3% (n=31) of the respondents diagnosed with diabetes, were unable to recognise the disease.

### Sociodemographic factors associated with correct recognition

Logistic regression modelling was used to determine the significant correlates of the recognition of diabetes. Table 3 revealed that individuals of Malay ethnicity (odds ratio [OR], 1.39, 95% confidence interval [CI], 1.02 to 1.89; P= 0.04) had higher odds of correct recognition for diabetes than the Chinese. Compared to individuals of ages 18 to 34 years, the recognition of diabetes was significantly higher among the age groups of 35 to 49 years (OR, 1.85; 95% CI, 1.15 to 2.98; P= 0.01) and 50 to 64 years (OR, 2.06; 95% CI, 1.19 to 3.56; P= 0.01). Individuals who were diagnosed with diabetes were nearly three times as likely to correctly recognise the condition as those who did not have diabetes (OR, 2.64; 95% CI, 1.38 to 5.08; P < 0.01).

**Table 3.** Sociodemographic correlates of correct recognition of diabetes.

|  | OR | 95% CI |  | P value |
| --- | --- | --- | --- | --- |
| Age group |  |  |  |  |
| 18 to 34 | Ref |  |  |  |
| 35 to 49 | 1.85 | 1.15 | 2.98 | 0.01 |
| 50 to 64 | 2.06 | 1.19 | 3.56 | 0.01 |
| 65 and above | 1.45 | 0.76 | 2.75 | 0.26 |
| Gender |  |  |  |  |
| Female | Ref |  |  |  |
| Male | 0.64 | 0.46 | 0.90 | 0.01 |
| Ethnicity |  |  |  |  |
| Chinese | Ref |  |  |  |
| Malay | 1.39 | 1.02 | 1.90 | 0.04 |
| Indian | 1.24 | 0.92 | 1.68 | 0.15 |
| Others | 0.59 | 0.37 | 0.93 | 0.02 |
| Education |  |  |  |  |
| Primary and below | 0.58 | 0.31 | 1.11 | 0.10 |
| Secondary | 0.75 | 0.43 | 1.32 | 0.32 |
| Pre-University/Junior College | 0.79 | 0.36 | 1.71 | 0.54 |
| Vocational Institute/ITE | 0.48 | 0.26 | 0.89 | 0.02 |
| Diploma | 0.91 | 0.54 | 1.53 | 0.73 |
| Degree and above | Ref |  |  |  |
| Marital status |  |  |  |  |
| Married/Cohabiting | Ref |  |  |  |
| Single | 1.04 | 0.67 | 1.60 | 0.87 |
| Divorced/separated | 0.93 | 0.45 | 1.92 | 0.85 |
| Widowed | 0.85 | 0.40 | 1.80 | 0.67 |
| Employment |  |  |  |  |
| Employed | Ref |  |  |  |
| Economically Inactive<sup>a</sup> | 1.07 | 0.69 | 1.66 | 0.77 |
| Unemployed | 0.48 | 0.25 | 0.92 | 0.03 |
| Monthly Income (SGD) |  |  |  |  |
| Below 2,000 | Ref |  |  |  |
| 2,000 to 3,999 | 1.21 | 0.78 | 1.88 | 0.39 |
| 4,000 to 5,999 | 1.22 | 0.66 | 2.28 | 0.53 |
| 6,000 to 9,999 | 1.50 | 0.68 | 3.30 | 0.32 |
| 10,000 and above | 1.25 | 0.51 | 3.06 | 0.62 |
| No income | 0.98 | 0.53 | 1.81 | 0.95 |
| Diabetes status |  |  |  |  |
| No | Ref |  |  |  |
| Yes | 2.64 | 1.38 | 5.08 | 0.004 |

<sup>a</sup>Economically inactive includes retired, homemaker, student, and the physically disabled; SGD: Singapore Dollar

However, we found that male gender (OR, 0.64; 95% CI, 0.46 to 0.90; P < 0.01) and those from the Others ethnic category (OR, 0.59; 95% CI, 0.37 to 0.93; P < 0.01), had significantly lower odds of recognition for diabetes. Those who were unemployed (OR, 0.48; 95% CI, 0.25 to 0.92; P= 0.03) also had lower odds of correct recognition compared to those who were employed.

## DISCUSSION

This was the first extensive study of diabetes literacy in in Singapore to evaluate recognition by using a vignette. The study also identified the significant sociodemographic characteristics associated with the correct recognition of diabetes mellitus. This study serves as a baseline measure of diabetes knowledge in Singapore’s general population and will play an important role to inform future health policies and initiatives as part of ongoing national efforts to tackle diabetes.

Overall, the findings of this study suggest that the Singaporean adults have a relatively high rate (82.7%) of diabetes recognition. Our results present a striking contrast with other research [15] and much higher than previously found local estimates, where only about 60% of the general population were able to recognise symptoms and complications of diabetes.[17] In a vignette-based study by Vimalanathan and Furnham,[22] which explored health literacy of different types of diseases, diabetes had a correct recognition rate of 46% among the British adults. Diabetes literacy in our study, in this regard, was considerably higher. The high percentage of Singaporeans recognising diabetes may be a positive reflection of the recent concerted national efforts and developments surrounding diabetes awareness in the country, which has led to a slew of measures including public health campaigns (‘War on Diabetes’), education initiatives, and the portrayal of diabetes-related problems and complications in local mass media.[23] However, some individuals though aware of this disease, may struggle to label it in proper medical terms. This was reflected in our results, with a proportion of answers observed to be “partly correct recognition”, wherein diabetes was labelled in non-technical, colloquial language (e.g., “high blood sugar”). Another interesting finding includes some individuals interpreting the symptoms depicted in the vignette as the presence of other medical conditions. This was reflected in our analysis with 7.7% of the respondents ascribing it to health problems other than diabetes. For example, symptoms of “feeling thirsty” were construed to be a condition of “dehydration”. This suggests people may be using their general health knowledge to answer, rather than having specific knowledge about diabetes. While we can appreciate that recognising symptoms as any other physical health condition (not diabetes) might still prove helpful in prompting individuals to seek professional care, ascribing the symptoms to a less serious condition can lead to significant delays in seeking proper and effective treatment. Furthermore, this points out a gap in the public knowledge of diabetes which must be addressed in order to better differentiate its symptoms and treatments, from other health problems. The signs and symptoms of diabetes are overlooked because of the slow and chronic progression of the disease; unlike many other physical conditions, the consequences of diabetes may not be manifested immediately.[24]

The current study identified a number of sociodemographic factors significantly associated with correct recognition, including age, gender, ethnicity, as well as personal experience with diabetes. Recognition varied by age group, where young individuals (18 to 34 years). recognised diabetes most poorly, whereas those aged between 35 to 49 years, and 50 to 64 years were significantly better at correctly recognising diabetes. We surmise that these findings may have been a result of several factors. Firstly, the average age of onset of diabetes is about 45 years.[4] Comparably, the average age of respondents in our study was 45 years and a substantial proportion of adults with self-reported diabetes were from the age groups of 35-49 and 50-64 who may have experience in dealing with similar problems to those described in the vignette which aids better recognition. Unlike mental illnesses which may present with an early onset in younger people,[25] chronic physical diseases are known to be largely present and of greater concern in older individuals.[26] Perhaps the young adults in this study, were less likely to have directly or indirectly encountered diabetes. This theory is consistent with the local health report[7] which suggests young adults were less likely to have attended health screenings for chronic disease and were less likely to seek frequent treatment or follow up consultation for diabetes than individuals from older age groups. Therefore, it is not surprising that the knowledge and ability to correctly recognise signs and symptoms of diabetes among young adults is poorer than those of older ages. These findings add to the importance of re-thinking the diabetes public education efforts in Singapore. There may be a need for different educational strategies, focusing on specific symptoms of the disease, and a consideration of how the content is communicated, so as to better target young adults given recognition of diabetes was poorest among them.

This study also observed a significant difference between males and females’ ability to recognise the physical disease. There are in diverse populations Consistent with the body of literature which reports better rates of diabetes recognition among females than males,[5, 27] our study found that males were specifically less able to recognise diabetes. Gender differences in both mental and physical health literacy have previously been explained by greater self-awareness and higher sensitivity to symptoms of illnesses among women compared to men.[28, 29] This gender disparity has also been associated with health behaviour paradigms such that, men are less likely to seek professional help for their health problems [30-32] which could contribute to men’s health gap in terms of disease knowledge and recognition. Therefore, there is a crucial need for policy and health promotion programs to target males who have a higher burden of the disease, with diabetes education strategies and materials designed to suit their needs and characteristics.

In this study, we found a relationship between ethnicity and recognition of diabetes. That is, ethnic Malays were more likely to identify it correctly, and those from the ethnic group, Others, were less likely to correctly recognise diabetes compared to Chinese. It is possible that respondents who are ethnic Malay, were disproportionately living with diabetes compared with the Chinese majority [33] or more likely to have direct contact with other individuals living with the disease. It can also be reasoned that the national diabetes education campaign since 2016, meant to engage ethnic Malays through community programs and initiatives, may have been successful.[34] This emphasises utility in encouraging community involvement to support diabetes education by engaging racial/religious community groups in several small-scale activities. On the other hand, individuals from the Others ethnic group demonstrated poor recognition of diabetes, which could be due to the attribution of the symptoms to causes other than the disease. This is unfortunate because recognition of early symptoms can help to get the disease under control immediately and prevent long-term complications. Content analysis of incorrect responses showed that there was a general tendency to consider symptoms described in the vignette as ‘not a medical problem’ or misinterpreting symptoms as ‘ageing’ or ‘other medical problems’. However, we are unable to offer any definite explanation for this phenomenon just as has been observed in other research.[35] Future studies to elucidate underlying reasons for ethnic differences is warranted.

Employment status was another significant predictor of diabetes knowledge. Compared to those employed, unemployed individuals were found to be less likely to correctly recognise diabetes in our study. The majority of diabetes cases occur among working adults.[36] Research has linked diabetes to physical disability in adults worldwide,[37, 38] resulting in increased sick days for employees and increased costs for employers.[39] Thus, workplace screening and health talks for diabetes may lead to greater awareness and better recognition among those who are employed. In Singapore, health talks and routine health screenings including comprehensive blood tests offered to employees have become increasingly commonplace.[40] There may be easier affordability to utilise healthcare services among residents who are gainfully employed compared to their unemployed peers. This is consistent with a recent national survey which found that participation in health screenings rose in tandem with income.[7] It was not unexpected that persons with diabetes had significantly better recognition of the disease than the healthy population in our study. Patients with diabetes would have a deeper personal understanding of symptoms and chronic complications of diabetes when compared to the healthy population given their own experience and the education imparted as part of management.

### Limitations

There are several limitations. The vignette format might have facilitated diabetes identification by clearly describing persons with prototypical symptoms and functional changes. Recognition may be poorer in real life, because one might not notice the slow changes and ignore, minimise, or misattribute symptoms to other causes. The recognition rate found here may not apply to non-typical presentations of diabetes.[41] Moreover, the study did not include those who had language difficulties, and those who were institutionalised, hospitalised, or uncontactable during the survey period. Our results could have overestimated diabetes recognition in the population. Nevertheless, the current study has its strengths in that it was a nationwide study with a representative sample who were surveyed ensuring high quality of the data collected. Additionally, the vignette was developed with inputs from experienced clinicians who are experts in the field and further cognitive testing was undertaken before use. Also, the study instruments were translated into the three major local languages ensuring inclusivity.

## Conclusion

Given the high prevalence rate of diabetes in high-income countries including Singapore, it is imperative that educational campaigns target all members of the population with accurate and appropriate information. In order to do this, it is important to establish the population’s baseline knowledge and ability to recognise the signs and symptoms of diabetes. This study therefore set out to obtain up-to-date data, which future strategies and national programs could potentially deploy. Our findings can inform strategic plans to address the growing diabetes epidemic. To prevent diabetes, reduce its economic burden, and improve the quality of life for Singaporean adults with diabetes or at risk of diabetes, public health messages and healthcare system interventions should target specifically, young adults, males, and the unemployed with poor knowledge of diabetes. This should include the development of suitable strategies to communicate more effectively with a deeper understanding of the needs and competencies of the specific demographic. Research to develop effective ground-up community initiatives to more widely apply diabetes education programs should be supported.

## Data Availability

The data that support the findings of this study are available from the corresponding author upon reasonable request.

## Contributors

KR led the analysis plan and interpretation of findings and prepared the manuscript. MS and AP were actively involved in the analysis, interpretation and manuscript content. MS, SAC, ESL and CFS were involved in the conceptualisation of the study. JHL conducted the analysis and assisted in interpretation of findings and manuscript content. FD and PW provided significant intellectual inputs into the manuscript.

## Patient and public involvement

Cognitive testing of survey questionnaires involved patients with diabetes and members of the public to improve the quality of the questionnaire and to adapt it for local population use. Patients and/or the public were not involved in the recruitment, conduct, reporting and dissemination plans of this research.

## REFERENCES

1. World Health Organization. Global report on diabetes, 2016. Available: https://www.who.int/diabetes/global-report/en/ [Accessed 2 Dec 2019].

2. Cho NH, Shaw JE, Karuranga S, et al. IDF diabetes atlas: global estimates of diabetes prevalence for 2017 and projections for 2045.Diabetes Res Clin Pract 2018;138:271– 81

3. Baynes HW. Classification, pathophysiology, diagnosis and management of diabetes mellitus. J diabetes metab. 2015 May 1;6(5):1–9.

4. Png ME, Yoong J, Phan TP, et al. Current and future economic burden of diabetes among working-age adults in Asia: conservative estimates for Singapore from 2010-2050. BMC Public Health 2016;16:153

5. Ng CS, Toh MP, Ko Y, Lee JY. Direct medical cost of type 2 diabetes in Singapore. PloS One. 2015 Mar 27;10(3):e0122795.

6. Epidemiology & Disease Control Division. The burden of disease in Singapore, 1990–2017: an overview of the global burden of disease study 2017 results. Seattle, WA: IHME, 2019. Available: http://www.Healthdata.Org/sites/default/files/files/policy_report/2019/GBD_2017_Singapore_Report.pdf [Accessed 1 July 2020]

7. Ministry of Health. National Health Survey. 2011. Available: https://www.moh.gov.sg/content/dam/moh_web/Publications/Reports/2011/NHS2010%20-%20low%20res.pdf [Accessed 1 July 2020].

8. Jorm AF. Mental health literacy: public knowledge and beliefs about mental disorders. Br J Psychiatry 2000;177:396–401

9. Vijan S, Stevens DL, Herman WH, Funnell MM, Standiford CJ. Screening, prevention, counseling, and treatment for the complications of type II diabetes mellitus: putting evidence into practice. J Gen Intern Med. 1997 Sep;12(9):567–80.

10. Abdullah A, Liew SM, Ng CJ, Ambigapathy SV. Paranthaman PV. Health literacy experiences of multi-ethnic patients and their health-care providers in the management of type 2 diabetes in Malaysia: A qualitative study. Health Expect. 2020 Oct;23(5):1166–76.

11. Leung AYM, Bo A, Hsiao HY, Wang SS, Chi I. Health literacy issues in the care of Chinese American immigrants with diabetes: a qualitative study. BMJ Open. 2014;4(11):1–12

12. Prabsangob K, Somrongthong R, Kumar R. Health literacy among Thai elderly population with type-2 diabetes living in rural area of Thailand. Pak J Health. 2018;8(1):27–31

13. Soong FS. Beliefs and practices of Chinese diabetic patients concerning the cause and treatment of their ill-health. Singapore Med J. 1971 Dec 1;12(6):309–13.

14. Adelene C, Thelma C, Drishti S, Cesidio P, Ezekiel UN. Diabetes mellitus literacy in a regional community of a developed country. Acta Biomed. 2020;90(4):482.

15. Kayyali R, Slater N, Sahi A, Mepani D, Lalji K, Abdallah A. Type 2 Diabetes: how informed are the general public? A cross-sectional study investigating disease awareness and barriers to communicating knowledge in high-risk populations in London. BMC Public Health. 2019 Dec;19(1):1–1.

16. Herath HM, Weerasinghe NP, Dias H, Weerarathna TP. Knowledge, attitude and practice related to diabetes mellitus among the general public in Galle district in Southern Sri Lanka: a pilot study. BMC Public Health. 2017 Dec;17(1):1–7.

17. Wee HL, Ho HK, Li SC. Public awareness of diabetes mellitus in Singapore. Singapore Med J. 2002 Mar 1;43(3):128–34

18. Department of Statistics, Singapore. Census of Population, Population Trends. 2019. Available: https://www.singstat.gov.sg/-/media/files/publications/population/population2018.pdf [Accessed 25 July 2020]

19. AshaRani PV, Abdin E, Kumarasan R, et al. Study protocol for a nationwide Knowledge, Attitudes and Practices (KAP) survey on diabetes in Singapore’s general population. BMJ Open. 2020;10:e037125. doi:10.1136/bmjopen-2020-037125

20. Barter C, Renold E: ‘I wanna tell you a story’: exploring the application of vignettes in qualitative research with children and young people. Int J Soc Res Methodol. 2000, 3 (4): 307-323. 10.1080/13645570050178594

21. Hughes R: Using vignettes in qualitative research. Sociol Health Illn. 1998, 20(3):381-400. 10.1111/1467-9566.00107.

22. Vimalanathan A, Furnham A. Comparing physical and mental health literacy. J Ment Health. 2019 May 4;28(3):243–8.

23. Ministry of Health. War on diabetes. 2018. Available: https://www.moh.gov.sg/wodcj [Accessed 25 July 2020]

24. Ramachandran A. Know the signs and symptoms of diabetes. Indian J Med Res. 2014 Nov;140(5):579–81. PMID: 25579136; PMCID: PMC4311308

25. Kessler RC, Berglund P, Demler O, Jin R, Merikangas KR, Walters EE. Lifetime prevalence and age-of-onset distributions of DSM-IV disorders in the National Comorbidity Survey Replication. Arch Gen Psychiatry. 2005 Jun 1;62(6):593–602.

26. Chang J, Mosenifar Z. Differentiating COPD from asthma in clinical practice. J Intensive Care Med. 2007 Sep;22(5):300–9.

27. dos Santos PF, Dos Santos PR, Ferrari GS, Fonseca GA, Ferrari CK. Knowledge of diabetes mellitus: does gender make a difference? Osong Public Health Res Perspect. 2014 Aug 1;5(4):199–203

28. Bianchin M, Angrilli A. Gender differences in emotional responses: A psychophysiological study. Physiol Behav. 2012 Feb 28;105(4):925–32.

29. Van Wijk CG, van Vliet KP, Kolk AM, Everaerd WT. Symptom sensitivity and sex differences in physical morbidity: a review of health surveys in the United States and The Netherlands. Women Health. 1991;17(1):91–124.

30. Redondo-Sendino A, Guallar-Castillon P, Banegas JR, Rodriguez-Artalejo F. Gender differences in the utilization of health-care services among the older adult population of Spain. BMC Public Health 2006;6:155.

31. Jonsson PM, Sterky G, Gafvels C, Ostman J. Gender equity in health care: the case of Swedish diabetes care. Health Care Women Int. 2000;21(5):413–31

32. Chang HY, Hsu CC, Pan WH, Liu WL, Cheng JY, Tseng CH, Bai CH, Yeh WT, Hurng BS. Gender differences in trends in diabetes prevalence from 1993 to 2008 in Taiwan. Diabetes Res Clin Pract. 2010 Dec 1;90(3):358–64.

33. Phan TP, Alkema L, Tai ES, Tan KH, Yang Q, Lim WY, Teo YY, Cheng CY, Wang X, Wong TY, Chia KS. Forecasting the burden of type 2 diabetes in Singapore using a demographic epidemiological model of Singapore. BMJ Open Diabetes Res Care. 2014 Jun 1;2(1):e000012.

34. Campaign launched to help Malays fight diabetes. 2016. Available: https://www.todayonline.com/singapore/six-month-nationwide-campaign-be-rolled-out-ccs-fight-diabetes-oct [Accessed 4 January 2021].

35. Chinnappan S, Sivanandy P, Sagaran R, Molugulu N. Assessment of knowledge of diabetes mellitus in the urban areas of Klang district, Malaysia. Pharmacy. 2017 Mar;5(1):11.

36. Nixon H, Robertson D. The role of occupational health in diabetes management. Occup Health (Lond). 2008 Apr 1;60(4):29.

37. Szosland D. Shift work and metabolic syndrome, diabetes mellitus and ischaemic heart disease. Int J Occup Environ Med. 2010 Jul 1;23(3):287.

38. Wong E, Backholer K, Gearon E, Harding J, Freak-Poli R, Stevenson C, Peeters A. Diabetes and risk of physical disability in adults: a systematic review and meta-analysis. Lancet Diabetes Endocrinol. 2013 Oct 1;1(2):106–14.

39. Gulley T, Boggs D, Mullins R, Brock E. Diabetes screening in the workplace. Workplace Health Saf. 2014 Nov;62(11):444–6.

40. HPB Health at Work. Healthy Workplace Ecosystem. 2019. Available: https://www.hpb.gov.sg/workplace/healthy-workplace-ecosystem [Accessed 4 January 2021].

41. Sjöholm Å. Atypical diabetes: a diagnostic challenge. BMJ Open Diabetes Res Care. 2020 Aug 1;8(1):e00147

