## Appendix A for "Recognition of Diabetes in a Multi-ethnic Population: Results from a Nationally Representative Population-Based Study"

**Sample of the diabetes vignette used in this study.**

"Mr.Tan is a 68-year-old male who has found activities like oil painting to fill his days since retiring from his job three years ago. Mr.Tan keeps himself busy in the morning by looking after his plants in the corridor of his HDB apartment. He notices that he seems to be going to the bathroom (urinating) quite often. After eating lunch with his wife, Mr.Tan takes a walk around the block. He feels extremely tired and very thirsty upon returning home. He also notices that the wound on his foot is taking a long time to heal. He does not work on his painting sometimes because his vision is blurry."

**Gender and ethnicity of the character presented in the vignette.**

| Race | Male | Female |
|---|---|---|
| Chinese | Mr. Tan | Mdm Tan |
| Malay | Mr. Ali | Mdm Siti |
| Indian | Mr. Raam | Mdm Rani |
| Other | Mr. John | Mdm Mary |
